# Accelerated intermittent theta burst as a substitute for patients needing electroconvulsive therapy during the COVID-19 pandemic: study protocol for an open-label clinical trial

**DOI:** 10.1101/2020.12.15.20248260

**Authors:** Daniel M. Blumberger, Zafiris J. Daskalakis, Fidel Vila-Rodriguez, David Boivin-Lafleur, Michelle S. Goodman, Tyler Kaster, Yuliya Knyahnytska, Gerasimos Konstantinou, Alisson P. Trevizol, Daphne Voineskos, Cory R. Weissman, Jonathan Downar

**Author notes:** Corresponding author: Dr. Daniel M. Blumberger, 1001 Queen St. W., Unit 4-115, Toronto, ON, Canada M6J1H4.

## Abstract

**Background:** Treatment resistant depression (TRD) is one of the leading causes of disability in Canada and is associated with significant societal costs. Repetitive transcranial magnetic stimulation (rTMS) is an approved, safe, and well-tolerated intervention for TRD. In the setting of the COVID-19 pandemic, reducing the number of visits to the clinic is a potential approach to significantly minimize exposure and transmission risks to patients. This can be accomplished by administering multiple treatment sessions in a single day, using an rTMS protocol known as accelerated intermittent theta burst stimulation (aiTBS). The objective of this novel study is to assess the feasibility, acceptance and clinical outcomes of a practical high-dose aiTBS protocol, including tapering treatments and symptom-based relapse prevention treatments, in patients with unipolar depression previously responsive to electroconvulsive therapy (ECT) or patients warranting ECT due to symptom severity.

**Methods:** All patients with unipolar depression referred to the brain stimulation service at the Centre for Addiction and Mental Health (CAMH) who warrant ECT will be offered screening to assess for eligibility to enroll in this trial. This open label, single group trial consists of 3 phases. In the acute treatment phase, treatment will occur 8 times daily for 5 days a week, until symptom remission is achieved or a maximum of 10 days of treatment. In the tapering phase, treatments will be reduced to 2 treatment days per week for 2 weeks, followed by 1 treatment day per week for 2 weeks. Patients will then enter the symptom-based relapse prevention phase including virtual check-ins and a treatment schedule based on symptom level. Remission, response and change in scores on several clinical measures from baseline to the end of the acute, tapering and relapse prevention phases represent the clinical outcomes of interest.

**Discussion:** Findings from this novel clinical trial may provide support for the use of aiTBS, including tapering treatments and symptom-based relapse prevention treatments, as a safe and effective alternative intervention for patients needing ECT during the COVID-19 pandemic.

**Trial registration:** Clinicaltrials.gov: NCT04384965

## Introduction

### Background, rationale and objectives

Major depressive disorder (MDD) is a highly prevalent and disabling disorder associated with substantial societal costs (1). Among Neurological, Mental, and Substance Use disorders, MDD is the number one cause of disability worldwide (2). In Ontario, Canada alone, $12.5 billion a year is attributed to mood disorder-related costs (3). The magnitude of the social, health, and economic burden of MDD reflects, to a large degree, the limited effectiveness of our current treatment options. Treatment-resistant depression (TRD) is defined as failure to respond to at least 2 adequate antidepressant trials (4) – a distressingly common problem. Electroconvulsive therapy (ECT) achieves remission rates of 65-75% in TRD (5, 6), but is hampered by significant cognitive adverse effects (7-9), public stigma (10), and the need for anesthesia, limiting its use to <1% of TRD cases (11). In the setting of the COVID-19 pandemic, access to ECT will likely be even lower due to restriction in volumes related to enhanced infection control procedures.

Repetitive transcranial magnetic stimulation (rTMS) is an evidence-based treatment for TRD. It involves stimulation of cortical neurons using externally applied, powerful, focused magnetic field pulses to induce lasting changes in the activity of brain regions involved in regulating thoughts, emotions, and behavior (12-14). Stimulation is applied non-invasively, over the scalp, using a handheld magnetic coil (15). Unlike with ECT, treatment does not require seizure induction, anesthesia, or the accompanying necessary manual ventilation that can aerosolize respiratory droplets, which is a major concern due to the COVID-19 pandemic.

rTMS is approved as a treatment for MDD by both Health Canada (2002) and the US Food and Drug Administration (FDA) (2008). The standard rTMS protocol for TRD, approved by the FDA, by far the most widely used across North America, involves applying 10 Hz stimulation to the left dorsolateral prefrontal cortex (DLPFC). Treatment consists of 3000 pulses delivered over a period of 37.5 minutes (16) and can last between four to six weeks, totaling 20 to 30 daily sessions per patient. In the setting of the COVID-19 pandemic, reducing the number of visits to the clinic is important to minimize exposure and transmission risk to patients. This can be accomplished by administering multiple treatment sessions on a single day, an approach known as accelerated rTMS. This protocol allows patients to receive a complete course of rTMS with fewer clinic visits (though each visit lasts a day) overall. In effect, this protocol cohorts patients into smaller groups receiving treatment at the same time.

Recently, our research teams achieved considerable gains in improving rTMS cost-effectiveness by reducing the time required per session and conducted the landmark trial that led to FDA approval of intermittent theta burst stimulation (iTBS) (17). As opposed to standard rTMS, iTBS uses 50 Hz bursts of stimulation and can be administered in 1/10^th^ of the time of high frequency rTMS (∼3 min vs. 37.5 min), yet achieves similar reductions in depressive symptom burden (18). An accelerated protocol with multiple daily brief iTBS sessions could enable patients to achieve remission more rapidly and reduce time of direct contact with others. Thus, the clinical potential of accelerated iTBS (aiTBS) protocols in patients who would otherwise get ECT is substantial.

Several recently published studies have demonstrated that accelerated rTMS can achieve similar outcomes to once daily rTMS treatment in only 4 to 10 days (19, 20). A recent study found that applying 2 sessions per day for 10 days in 28 participants resulted in 56% response and 37% remission (21). A similar study applied 5 sessions per day for 4 days and achieved 35% response and 15% remission in 20 participants (22). More recently, researchers published a case series demonstrating that a 5-day regimen of 10x daily iTBS sessions is not only safe and tolerable but yields substantially higher remission rates than conventional once-daily rTMS (i.e., >70% remission on all measures) (23). Notably, this case series included patients who had previously failed to respond to once daily rTMS. Another study by researchers at Stanford University used the same treatment regimen and included patients with highly refractory MDD who did not respond to traditional high frequency rTMS and failed to respond to an acute course of ECT. Five of six participants responded to treatment, and four participants were in full remission at the final treatment session (24). Of note, these protocols (23, 24) include functional Magnetic Resonance Imaging (fMRI) imaging and a complicated algorithm to refine the treatment target location. Such procedures using neuroimaging guidance are neither feasible nor practical during a pandemic.

Unfortunately, in both of the latter studies, the response to treatment was not consistently maintained. Cole and colleagues reported that 6 of 21 participants underwent a second course after they no longer met remission criteria (23), and Williams and colleagues reported that by 4 weeks after treatment end, participants no longer met response criteria (24). Taken together, these findings suggest that while accelerated iTBS (aiTBS) may be a safe, well-tolerated, and effective treatment option, there is a clear need for continued investigation into maintaining response and remission rates following the acute intervention. As such, the primary objective of this study is to assess the feasibility, acceptance, and clinical outcomes of a practical high-dose aiTBS protocol, including tapering treatments and symptom-based relapse prevention treatments, in patients with MDD either previously responsive to ECT or those needing ECT due to symptom severity.

## Methods

### Study Design

All patients referred for ECT at the Centre for Addiction and Mental Health (CAMH) in Toronto, Canada, will be assessed by one of the brain stimulation psychiatrists as part of current clinical procedures. Suitable patients will be referred for virtual screening via a hospital approved videoconferencing software by a research analyst (RA). Prior to the screening visit, participants will be provided a link to a read-only copy of the consent form. The consent discussion will be conducted virtually, or over the telephone if the participant declines videoconferencing. Following the consent discussion, research personnel will obtain the participant’s informed e-consent to participate in the trial, using REDCap. If a prospective participant does not have access to internet/email/technology, they may come in-person to CAMH to give consent virtually, using our tablet or computer, or a paper copy can be mailed to them.

Weekly follow-up assessments, including clinical rating scale assessments and self-report questionnaires, will be conducted virtually via videoconferencing with a physician backup to the RA available at all times.

**Figure 1.**
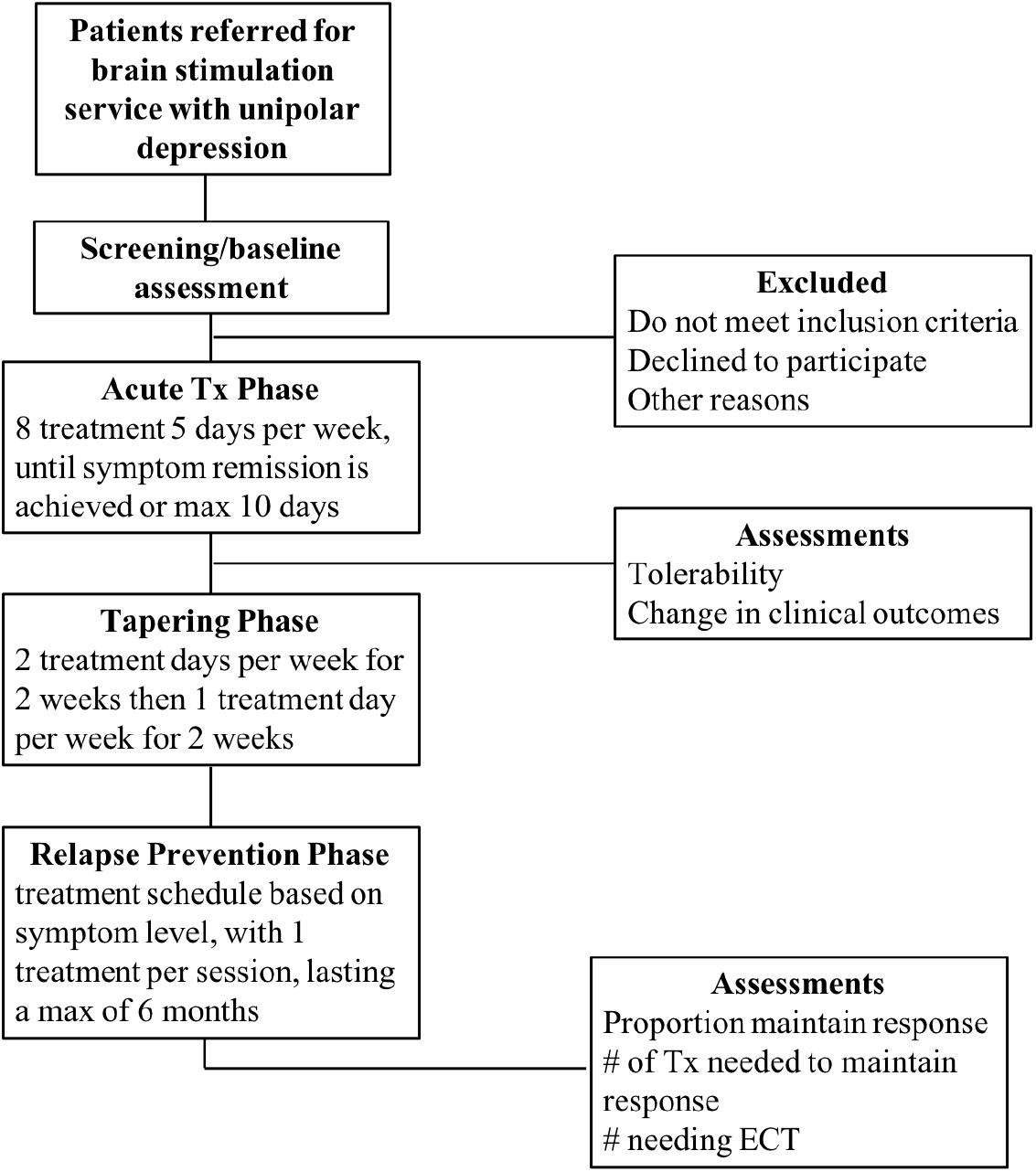
Flow diagram for the study design.

In the acute treatment phase, treatment will occur 8 times daily (50 min pause between treatments) on weekdays, until symptom remission is achieved (Hamilton Rating Scale for Depression, 24 item (HRSD-24) score ≤ to 10) or a maximum of 10 working days of daily treatment. In the tapering phase, treatments will be reduced to 2 days per week for 2 weeks and then 1 day per week for 2 weeks (4 weeks total). Patients who have an insufficient response after a minimum of 5 treatment days and maximum of 16 treatment days (i.e. the end of tapering phase) based on less than a 50% reduction in HRSD-24 from the screening visit and the clinical opinion of the brain stimulation psychiatrist, will be discontinued from the trial. Patients who have responded (minimum of 50% reduction in HRSD-24 scores) to treatment will enter the symptom-based relapse prevention phase including virtual check-in with the RA and a treatment schedule based on symptom level and clinician judgement. This modified relapse prevention algorithm, known as the STABLE algorithm, was developed to prevent relapse after a successful course of ECT (25). The relapse prevention phase will last a maximum of 6 months, consistent with prior ECT relapse prevention studies, and will include rescue treatment days (8 sessions per day) based on symptom re-emergence and clinician judgment. (see Table 1). Each treatment session across all phases of the study will consist of a single iTBS treatment, delivering 600 pulses of iTBS (bursts of 3 pulses at 50 Hz, bursts repeated at 5 Hz, with a duty cycle of 2 seconds on, 8 seconds off, over 20 cycles / ∼3 minutes) at a target of 110% of the subject’s resting motor threshold (RMT). This protocol involves minimal contact (3 min) between technician and patients.

**Table 1.** Study Phases and Relapse Prevention Algorithm

| Weeks 1-2: Acute Treatment Phase |  |  |  |
| --- | --- | --- | --- |
| Daily treatment (8 tx/day) for up to 10 days |  |  |  |
| Weeks 3-6: Tapering Treatment Phase |  |  |  |
| Weeks 3-4 |  | 2 treatment days (8 tx/day) per week for 2 weeks |  |
| Weeks 5-6 |  | 1 treatment day (8 tx/day) per week for 2 weeks |  |
| Weeks 6-30: Symptom-Driven Relapse Prevention Algorithm Treatment Phase |  |  |  |
| Treatment days per week | Description | Corresponding HRSD-24 ratings | Relapse Potential |
| 0 | Current symptom level very low | Current HRSD-24 score $\leq 6$ | Low |
| | Current symptom level low to moderate, with only small drift from baseline level | Current HRSD-24 score 7 - 12 and $\leq 2$ points higher than baseline | |
| | Last two HRSD-24 scores in remitted range with a flat trajectory (remission stable with less than 2-point change from previous) | Current HRSD-24 score 7 - 10 and previous score 5 -10 and current score is $\leq 2$ points higher than previous score or score is $\leq 2$ points higher than end of tapering score if not a remitter | |
| 2 | Current symptom level very high | Current HRSD-24 score $\geq 16$ | High |
| | Current symptom level moderate to high, with trajectory increasing rapidly and large drift from baseline | Current HRSD-24 score 11 -15, and current score is $\geq 3$ points higher than previous score, and current score is $\geq 8$ points higher than end of tapering score | |
| 1 | Patients not requiring 0 or 2 treatments receive 1 treatment | Current HRSD-24 score intermediate between criteria for low or high relapse potential | Moderate |

Technicians will follow local infection prevention and control (IPAC) recommendations regarding personal protective equipment and room cleaning. rTMS treatments will be conducted in person with added safety and cleaning precautions. Research staff will be wearing a mask, face shield, and gloves during any in-person interactions and will maintain a physical distance from patients whenever possible. In addition, all individuals will undergo screening for COVID-19 symptoms and contacts and will be provided with a mask, prior to entering CAMH each day. Any person who is found to have symptoms or contacts as per the screening protocol (known as a positive screen) will not be permitted to enter CAMH and treatment will not be allowed to continue that day. These cases will be reviewed by IPAC at CAMH to inform if, and when, treatment can continue.

### Clinical Measures

#### Screening evaluation

At the screening visit with the RA, patients will complete the Mini-International Neuropsychiatric Interview (MINI). This scale assesses current and lifetime MDD and other psychiatric disorders and it will be used to clarify psychiatric inclusion and exclusion criteria. The Antidepressant Treatment History Form (ATHF) will be used to collect information on previous anti-depressant treatment resistance. A detailed history of any prior ECT and rTMS treatment will be collected. The Transcranial Magnetic Stimulation Adult Safety Screen (TASS) will be used to assess potential rTMS safety risk factors. Patients will also complete baseline clinical measures including Hamilton Rating Scale for Depression (HRSD-24), Beck Depression Inventory (BDI-II), the Patient Health Questionnaire (PHQ-9), the Scale of Suicidal Ideation (SSI), the Generalized Anxiety Disorder 7-Item (GAD-7), and the World Health Organization Disability Assessment Schedule (WHODAS).

#### Clinical Assessments During Treatment

Depressive symptoms will be assessed using BDI-II (each treatment) and the HRSD-24 and the SSI (weekly). Hospital mandated assessments including the PHQ-9 and GAD-7 will be completed weekly and the WHODAS will be completed at the end of acute, tapering and relapse prevention phases. The clinician global impression severity (CGI-S) and improvement (CGI-I) scores will ve assessed by the most responsible physician. Remission, response and change in scores on the BDI-II, HRSD-24, GAD-7, PHQ-9 and SSI from baseline to the end of the acute, tapering and relapse prevention phases will be clinical outcomes of interest.

Regarding telephone assessments, two items on the HRSD-24 which pertain to the visual assessment of psychomotor retardation and agitation (i.e. Items 8 and 9) are not able to be completed. Therefore, these items will be scored as “0” and the clinical rater will document that the above items could not be rated. For self-report questionnaires completed at virtual assessment visits, the study personnel will complete the questionnaires with participants by: (1) reading the questions and answers verbatim to the participant and documenting their answers, (2) virtually sharing their screen so that the participant can read the questions and provide the answers, or (3) the research staff will send a link via REDCap to the participant to fill out the questionnaires on their own device.

If a participant reports worsening of symptoms or suicidal ideation or report adverse events that may require further investigation due to safety reasons, the study investigator or delegate will contact the participant for follow-up. If a participant reports feeling unsafe due to suicidal thoughts or requires medical attention, study personnel will advise to contact 911 or go to the nearest Emergency Room. All adverse events will be assessed and reported accordingly. If a participant unexpectedly drops off a virtual session, they will be called immediately to ensure there is no safety concern. The emergency contact will be contacted in case of emergency or an inability to contact the participant after they unexpectedly drop off a virtual session.

**Table 2:** Schedule of Events

| Measure | Screening<br>(Baseline) | Study Period |  |  |  |  |  |  |  |  |
| --- | --- | --- | --- | --- | --- | --- | --- | --- | --- | --- |
|  |  | Acute Tx Phase |  |  | Tapering Tx Phases |  |  | Relapse Prevention Tx Phase |  |  |
|  |  | Every Tx | Weekly/<br>Discontinuation | End/<br>Discontinuation | Every Tx | Weekly/<br>Discontinuation | End/<br>Discontinuation | Every Tx | Weekly/<br>Discontinuation | End/<br>Discontinuation |
| **Informed Consent** | X |  |  |  |  |  |  |  |  |  |
| **MINI** | X |  |  |  |  |  |  |  |  |  |
| **TASS** | X |  |  |  |  |  |  |  |  |  |
| **ATHF** | X |  |  |  |  |  |  |  |  |  |
| **ECT History** | X |  |  |  |  |  |  |  |  |  |
| **BDI-II** | X | X |  |  | X |  |  | X | X |  |
| **HRSD-24** | X |  | X |  |  | X |  |  | X |  |
| **PHQ-9** | X |  | X |  |  | X |  |  | X |  |
| **SSI** | X |  | X |  |  | X |  |  | X |  |
| **WHODAS** | X |  |  | X |  |  | X |  |  | X |

Tx: Treatment; MINI: Mini-International Neuropsychiatric Interview; TASS: Transcranial Magnetic Stimulation Adult Safety Screen; ATHF: Antidepressant Treatment History Form; BDI-II: Beck Depression Inventory; HRSD-24: Hamilton Rating Scale for Depression; PHQ-9: Patient Health Questionnaire; SSI: Scale of Suicidal Ideation

### rTMS Treatment Parameters

rTMS treatments will employ the MagPro X100 stimulator (MagVenture, Farum, Denmark) equipped with a B70 fluid-cooled coil. Prior to the first treatment (no more than 5 days prior), each subject’s motor threshold (MT) will first be determined according to published methods (26, 27). This location, as well as the stimulation target spot, will be marked at the first session on the scalp and standard methods will be used to target this spot during treatment sessions. The modified BeamF3 scalp heuristic will be used to localize the treatment site over the left DLPFC (28). A treatment day will consist of 8 treatment sessions, with the start of each session timed to be at least 50 minutes from the previous session.

#### Known Risks and Side Effects

Tens of thousands of people have received rTMS treatment over the last 20 years. As with any medical procedure, rTMS has certain risks, some of which are known. The known risks associated with rTMS are as follows (percentage of patients experiencing side effect):

- Common: Headache (30%), discomfort or pain at the stimulation site (20%), light headedness or dizziness after the treatment (20%), facial muscle twitching (30%). These side effects are mild and generally diminish over the course of treatment, and can usually be managed with rest, or with over-the-counter pain medications such as acetaminophen or ibuprofen.
- Less Common: (1-7%) fatigue, headache persisting after the treatment, dizziness or fainting during the initial sessions of rTMS treatment.
- Rare but Serious: Onset of suicidal thinking (less than 1%). There is also a 1% chance that rTMS may lead to a hypomanic episode (the opposite of depression, with symptoms elevated mood and energy, increased activity, impulsiveness, and decreased need for sleep). Participants will be monitored regularly for these symptoms during treatment and will have immediate access to a study psychiatrist if they appear.
- Very Rare but Serious: There are rare cases of an epileptic seizure resulting from rTMS (less than 0.1%). Safety guidelines have been in place since 1997 to minimize the risk of seizures from rTMS, and this study follows those guidelines. Still, worldwide, there have been four reports of a seizure during rTMS even when the guidelines were followed. In many of these cases, patients were taking medications known to increase the chances of a seizure occurring spontaneously (29).

Common side effects of rTMS treatment are expected and will be recorded separately from adverse events. If feasible, patients will be asked to defer any medication changes for 4 weeks prior to the treatment course. We will strongly discourage psychotropic medication initiation or increases other than medication to manage symptomatic burden (i.e. Trazodone or Zopiclone) during the course of rTMS to avoid confounding effects. A visual analog scale (0 no pain – 10 worst pain they have experienced) will be used for patients to rate the severity of pain from side effects as well as one open ended question asking about other side effects at every visit and after every treatment session. These questions will be completed by the technician so that side effect severity and resolution can be assessed and verified appropriately over time.

### Attendance and Withdrawal Criteria

Patients will be encouraged to attend all scheduled treatments. Those who meet the following criteria will be excluded from the per protocol analysis: If they (1) miss / fail to attend any one of the scheduled treatment days in the course overall; (2) miss / fail to attend more than 5 individual treatment sessions overall; (3) cannot tolerate stimulation at a minimum of 90% RMT for the entire session on more than 5 treatment sessions overall, as this is likely to lead to inadequate stimulation of the cortex; (4) experience worsening of symptoms warranting an immediate switch to ECT or another intervention, in the clinical opinion of a brain stimulation psychiatrist; (5) withdraw consent to participate; (6) have insufficient response after a minimum of 5 days of treatment and maximum of 16 treatments (i.e. the end of tapering treatments) based on less than 50% reduction in HRSD-24 or the clinical opinion of the brain stimulation psychiatrist; (7) experience sustained relapse during the relapse prevention phase, as defined by a need for more than two consecutive weeks of twice weekly treatment days. If a participant misses treatment(s) due to a failed COVID-19 screening and are not allowed to enter CAMH or receive treatment, these missed treatment(s) will not count towards discontinuation criteria. In such cases, the study investigators will decide whether treatment will be discontinued.

### Eligibility Criteria

#### Patients will be included if they

(1) are currently experiencing a unipolar depressive episode based on the MINI with or without psychotic symptoms; (2) have previous response to ECT or high symptom severity warranting acute ECT in the opinion of the brain stimulation psychiatrist; (3) are over the age of 18; (4) pass the TMS adult safety screening (TASS) questionnaire; (5) are voluntary and competent to consent to treatment.

#### Patients are excluded if they

(1) have a Mini-International Neuropsychiatric Interview (MINI) confirmed diagnosis of substance dependence or abuse within the last 1 month; (2) have a concomitant major unstable medical illness, cardiac pacemaker or implanted medication pump; (3) have a lifetime Mini-International Neuropsychiatric Interview (MINI) diagnosis of bipolar I or II disorder, schizophrenia, schizoaffective disorder, schizophreniform disorder, delusional disorder; (4) have any significant neurological disorder or insult including, but not limited to: a. any condition likely to be associated with increased intracranial pressure; b. space occupying brain lesion; c. any history of seizure except those therapeutically induced by ECT or a febrile seizure of infancy or single seizure related to a known drug related event; d. cerebral aneurysm; e. significant head trauma with loss of consciousness for greater than 5 minutes; (5) have an intracranial implant (e.g., aneurysm clips, shunts, stimulators, cochlear implants, or electrodes) or any other metal object within or near the head, excluding the mouth, that cannot be safely removed; (6) currently take more than lorazepam 2 mg daily (or equivalent) or any dose of an anticonvulsant due to the potential to limit rTMS efficacy; (7) lack of response to accelerated course of iTBS or rTMS in the past.

### Sample Size

All patients, including both in- and out-patients, referred to the ECT service with MDD with or without psychotic features who are deemed in need of ECT by one of the brain stimulation psychiatrists will be offered information on the trial and will be recorded. If patients do not consent to screening this will be recorded. No personal health information regarding patients who do not consent for screening will be collected. Collecting the number of referrals will allow for an assessment of the impact of the intervention on the entire service as a whole. We estimate that up to 200 patients will be treated with this protocol depending on the duration of the pandemic and the infection control measures instituted.

## Data Analysis and Management

### Clinical Outcomes Analysis

We will assess the number of patients with MDD referred to the service during the pandemic as described above. We will report the number that were able to complete the acute portion of the treatment schedule (up to 10 days) on both an outpatient and inpatient basis as a measure of feasibility. Tolerability will be assessed by reporting pain scores and number of withdrawals due to pain or other treatment related adverse events. Clinical outcomes will be described using descriptive statistics from baseline to post-treatment (paired t-test for continuous and McNemar’s chi-square for categorical variables) and will include change in clinical scales. The primary clinical outcome will be the proportion of patients achieving remission on the HRSD-24 (final score ≤ 10) between baseline and end of acute treatment and will be conducted on an intention to treat (ITT) and per protocol basis. Secondary clinical outcomes will include response on the HRSD-24 (≥ 50% reduction in scores) and change from baseline to end of acute treatment. We will repeat these analyses in the same manner for the other clinical scales used. Remission will be defined as scores ≤ 12 on BDI-II, < 5 on the GAD-7, ≤ 4 on the PHQ-9, and SSI = 0 (if score at baseline is ≥ 1). The change in WHODAS from baseline to end of acute and relapse prevention will also be reported. The proportion of patients maintaining response during relapse prevention will be reported along with number of treatments needed to maintain this response. In addition, the number needing to go on to receive ECT will be reported. Exploratory analyses will include evaluation of demographic and clinical variables associated with response.

### Data Management

The confidentiality of data collected and identity of the individuals participating in this study will be strictly maintained. All files pertaining to subjects in the study will be coded numerically and case report forms (CRFs) will not contain any personal health information. Source documents will be kept in a locked filing cabinet to limit access, and in the case of electronic source documents, files will be password-protected and saved in a secure server. REDCap software will be used for data collection and overall study data management over the course of this project. REDCap is an open-source, web-based clinical data management and electronic data capture system and database and was developed and managed in compliance with HIPAA, PIPEDA, and FDA 21 CFR Part 11 regulations. This system will be used to design electronic case report forms (eCRFs), data entry, data monitoring and cleaning, and for the query and export of datasets for statistical analysis and predictive modeling.

## Discussion

In summary, current treatment options for patients with TRD suffer from critical deficiencies of efficacy, capacity, or tolerability, especially given the current COVID-19 pandemic. rTMS and aiTBS, in particular, may be able to overcome many of these deficiencies as an approved, safe, well-tolerated, and effective treatment approach. The novel aiTBS protocol described here has the potential to rapidly help patients who otherwise would not be able to or may not want to receive ECT due to concerns regarding cognitive side effects, difficulties with accessibility or time commitment. Further, implementing aiTBS as a treatment for TRD may ultimately reduce the overall burden and need for ECT services, as demand for ECT treatment may exceed capacity during the COVID-19 pandemic. Importantly, shifting an rTMS service to deliver accelerated treatments reduces the number of patients coming into the clinic on a given day, ultimately minimizing exposure and transmission risks to patients. As such, we propose to study aiTBS, including tapering treatments and symptom-based relapse prevention treatments, as a substitute for patients needing ECT during the COVID-19 pandemic.

While several recently published studies have demonstrated that accelerated rTMS can achieve similar outcomes to once daily treatment, in a much shorter timeframe (19, 20, 23, 24), these studies also highlighted the risk of relapse following an acute course of aiTBS. The current study directly addresses the concerns regarding maintaining response and remission with the inclusion of the relapse prevention phase. In addition to potentially providing support for the use of aiTBS in maintaining response rates, we also aim to assess the feasibility, tolerability and clinical utility of aiTBS. An additional strength of this study is the predicted large sample sizes, given the volume of patients with MDD referred to the CAMH brain stimulation service.

The COVID-19 pandemic has resulted in unique challenges for both health care providers and researchers. Ultimately, this has led to a number of important modifications in how research is being conducted as well as several potential limitations. While a definitive assessment of aiTBS would require a randomized sham-controlled study, the ethical considerations of sham-controlled trials during this pandemic are more complicated than usual. Therefore, we have selected a single group, open-label trial study design.

Furthermore, unlike previous aiTBS protocols (23, 24) which include fMRI imaging to localize the treatment site, we will be using the modified BeamF3 scalp heuristic, given that neuroimaging is not feasible during a pandemic. We have also added a number of safety precautions, including virtual screening sessions and weekly assessments, daily symptom screening on site, and enhanced use of personal protective equipment and room cleaning. However, we will likely have to maintain a flexible study protocol to reflect the evolving infection prevention and control recommendations. Additionally, it is to be expected that COVID-19 may impact study recruitment and potentially increase the number of dropouts or missed treatments if patients screen or test positive. While the pandemic has complicated the way in which research is conducted, it is important to continue to adapt to keep rTMS services viable and perform it in the safest manner possible, both for patients and staff.

## Data Availability

The final dataset generated from the current protocol will be available from the corresponding author on reasonable request

## Trial status

The study is currently recruiting participants. Enrollment of participants started in May 2020.

## Abbreviations

aiTBS: accelerated intermittent theta burst stimulation
ATHF: Antidepressant Treatment History Form
BDI-II: Beck’s Depression Inventory
CAMH: Centre for Addiction and mental Health
CGI-S and CGI-I: clinician global impression severity and improvement
DLPFC: dorsolateral prefrontal cortex
eCRF: electronic case report forms
ECT: Electroconvulsive therapy
FDA: Food and Drug Administration
fMRI: functional Magnetic Resonance Imaging
GAD-7: Generalized Anxiety Disorder 7-Item
HRSD: Hamilton Rating Scale for Depression
ITT: intention to treat
MDD: Major depressive disorder
MINI: Mini-International Neuropsychiatric Interview
RMT: resting motor threshold
NMS: Neurological, Mental and Substance Use disorders
PHQ-9: Patient Health Questionnaire
RA: research analyst
rTMS: repetitive transcranial magnetic stimulation
SSI: Scale of Suicidal Ideation
STABLE: Symptom-Titrated Algorithm-based Longitudinal ECT
TASS: Transcranial Magnetic Stimulation Adult Safety Screen
TRD: treatment-resistant depression
WHODAS: World Health Organization Disability Assessment Schedule

## Authors’ contributions

DMB obtained funding and developed the manuscript, with important intellectual input from all co-authors.

## Ethics approval

Ethics approval was obtained by the Centre for Addiction and Mental Health Research Ethics Board (reference number 059/2020).

